# Intersex/Differences of Sex Development (I/DSD) Traits: Exploring Their Association with 15% of Phenotypically Detailed Genetic Disorders

**DOI:** 10.1101/2024.05.27.24307983

**Authors:** Leah Ragno, Louise C. Pyle

## Abstract

The study of Intersex traits or Differences of Sex Development (I/DSD) is an imperative area within medical genetics that confronts a long-standing problem: the prevalence of these conditions in the population is not fully clear, likely undercounted, and therefore no universally agreed upon numbers exist. Proportions among rare differences are not fully understood. While each specific I/DSD condition is individually rare, together they are estimated to affect about 0.7-1.7% of people worldwide (Fausto-Sterling, 1993). Over 10% of individuals with I/DSD traits exhibit comorbidities affecting other bodily systems, suggesting a broader, systemic impact of these conditions. The comprehensive phenotypic spectrum and systemic implications of I/DSD traits are not well-documented, likely due to social, cultural, and privacy concerns that may contribute to the under-phenotyping of genital variations. This research aims to delineate the disease landscape associated with I/DSD traits. By using the Online Mendelian Inheritance in Man (OMIM) database, a comprehensive resource for human genes and genetic disorders, we compiled an expert list of 155 Human Phenotype Ontology (HPO) terms that encompass the entirety of the medically defined I/DSD space. These terms were cross-referenced with the OMIM database to pinpoint diseases annotated with these HPO terms. Out of 8,181 phenotypically detailed diseases listed in OMIM and included in the HPO database, we discovered that 1,264 (15.5%) have associations with I/DSD-related HPO terms, signifying their presence in I/DSD traits. This substantial overlap underscores the need to rethink I/DSD not just as isolated anomalies but as a continuum of conditions that are crucial for a broader understanding of health issues. It also highlights that I/DSD traits are relatively common within the realm of rare differences, emphasizing the necessity for specialists, such as clinical geneticists, to be well-versed in I/DSD. Our findings call for a shift in the medical community’s perception and approach to studying I/DSD traits. Further research is required to confirm these associations and to explore the relationship between I/DSD traits and overall health.

## Introduction

Intersex traits or Differences of Sex Development (I/DSD) represent a complex and diverse group of conditions that challenge traditional binary notions of sex and gender. These conditions, which encompass a wide range of genetic, hormonal, and anatomical variations, are estimated to affect approximately 0.7-1.7% of the global population (Fausto-Sterling, 1993). Despite their prevalence, I/DSD conditions remain under-documented and poorly understood, partly due to social, cultural, and privacy concerns that hinder comprehensive phenotyping.

The intricate nature of I/DSD conditions often involves multiple organ systems, suggesting that these traits are not isolated anomalies but part of a broader spectrum of genetic disorders. Previous studies have indicated that over 10% of individuals with I/DSD traits exhibit comorbidities affecting other bodily systems, underscoring the systemic implications of these conditions (Cox et al., 2014). The interconnectedness of developmental pathways, particularly those involved in sex determination and gonad development, highlights the need for a holistic approach to understanding I/DSD traits.

This study aims to elucidate the disease landscape associated with I/DSD traits by leveraging the Online Mendelian Inheritance in Man (OMIM) database, a comprehensive resource for human genes and genetic disorders. We compiled an expert-curated list of 155 Human Phenotype Ontology (HPO) terms that encompass the medically defined I/DSD space. By cross-referencing these terms with the OMIM database, we identified diseases annotated with these HPO terms, revealing that 15.5% of the 8,181 phenotypically detailed diseases listed in OMIM have associations with I/DSD-related HPO terms.

Our findings underscore the need to reconceptualize I/DSD not merely as isolated conditions but as a continuum of traits with significant overlap across various genetic disorders. This perspective is crucial for enhancing the understanding of I/DSD traits and their broader implications for health. Furthermore, the study highlights the necessity for specialists, such as clinical geneticists, to be well-versed in the diverse presentations of I/DSD conditions.

## Methodology

The OBO, OWL, and JSON files (v2023-10-09) used in this integration were retrieved from The Human Phenotype Ontology’s primary web portal (Köhler et al., 2021).

### Assembly of an All-Encompassing I/DSD Diagnoses Index

We compiled an index of OMIM diseases and ORPHA syndromes, encompassing clinically agreed-upon I/DSD diagnoses. We identified these OMIM diseases and ORPHA syndromes from Differences of Sex Development and Ambiguous Genitalia disease panels, sourced from PreventionGenetics, a CLIA and ISO 15189:2012 accredited clinical DNA testing laboratory (Ambiguous genitalia panel.2024; Differences of sex development (DSD) panel.2024). To improve the comprehensiveness of our I/DSD trait index, we also identified all diagnoses associated with five (5) standard canonical I/DSD. These HPO terms were Ambiguous Genitalia (HP:0000062), Sex Reversal (HP:0012245), True Hermaphroditism (HP:0010459), Male Pseudo-hermaphroditism (HP:0000037), and Female Pseudo-hermaphroditism (HP:0010458). We then identified all HPO terms associated with the complete I/DSD trait index, from which to develop the I/DSD-Trait HPO Lexicon. The full index is available for review in Supplemental File 1.

### Formulation of I/DSD-related HPO Lexicons

We next crafted a lexicon of all HPO terms annotated to the assembled I/DSD traits index. This lexicon was then filtered to retain only those terms classified as ‘Genitourinary,’ resulting in a collection of 222 unique HPO terms. This collection, named the Comprehensive Genitourinary Lexicon for DSDs (CGL-DSD), was further refined by subject matter expert (L.C.P.) to exclude terms that were not medically defined I/DSD traits. The excluded HPO terms were primarily those pertaining to renal conditions. The resultant lexicon, termed the Focused Genitourinary DSD Glossary (FGD-Glossary), included 116 HPO terms that were highly specific and representative of I/DSD disorders. The comprehensive lists of HPO terms that constitute the CGL-DSD and the FGD-Glossary are in Supplemental File 2.

### Identifying Diseases Annotated to the FGD-Glossary

We then created a list of all OMIM diseases and ORPHA syndromes annotated to at least one HPO term in the FGD-Glossary. Our hypothesis was that if a disease was characterized by one of these HPO terms, it would provide valuable insights into its correlation with the broader scope and patterns of I/DSD disorders. Finally, we curated a list of all HPO terms annotated to this assemblage of FGD-Glossary-annotated OMIM diseases. These lists are available for review in *Supplemental File 3*.

### Statistical Analysis of HPO Annotations in FGD-Glossary-Annotated OMIM Diseases

To assess the distribution of inheritance patterns in FGD-Glossary-annotated OMIM diseases relative to the entire set of HPO-annotated OMIM diseases, we conducted a series of proportions z-tests. This statistical approach was selected to evaluate the significance of differences in the frequency of specific HPO Modes of Inheritance terms between the two groups. After computing the relative proportions of each term, we subjected these to the z-test and employed the Benjamini-Hochberg procedure to adjust the p-values and control the false discovery rate (FDR). Additionally, the same z-test was applied across all HPO annotations for the 1,264 FGD-Glossary-annotated OMIM diseases to ascertain which terms were significantly over- or underrepresented when compared to the entire HPO database OMIM dataset.

Finally, to assess the distribution of HPO annotations, we analyzed both individual terms and their broader categorizations into primary and secondary phenotypic categories, as defined by the hierarchy structured in the HPO OBO file (v2023-10-09) (Köhler et al., 2021). This hierarchy is depicted in **Figure 1**. This categorization allowed us to compare the frequency of HPO annotations within 1,264 FGD-Glossary-annotated OMIM diseases against the entire set of 8,181 OMIM diseases. We conducted a proportions z-test for each category to evaluate the presence of statistically significant differences in proportions. To address the issue of multiple comparisons, we once again applied the Benjamini-Hochberg procedure to adjust the resulting p-values and control the FDR. Further z-tests were then employed on selected primary and secondary categories that showed statistically significant (*p*-adj.<0.05) changes, allowing for a more detailed investigation into the specific areas of discrepancy in proportional representation between the two datasets.

**Figure 1:**
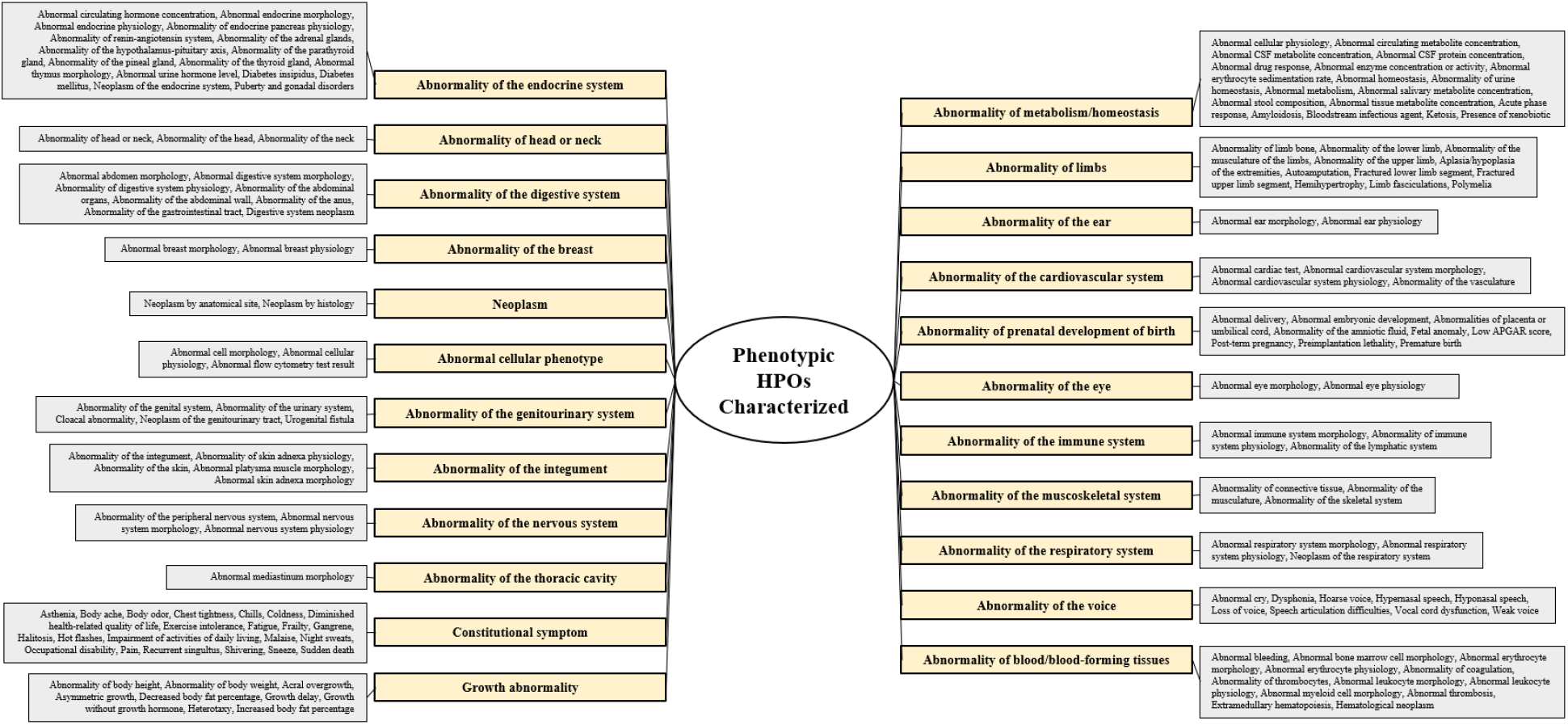
Hierarchical Structure of Phenotypic Characterizations in the HPO OBO File (v2023-10-09). This figure illustrates the categorization of individual HPO terms into primary and secondary phenotypic categories, as analyzed in the comparison of HPO annotations between FGD-Glossary-annotated OMIM diseases and the entire OMIM disease dataset.

Detailed categorizations and the full results of the statistical analyses are comprehensively presented in *Supplemental File 4*.

## Results

### Compilation of Representative I/DSD Disorders

Our methodology resulted in the identification of 362 disorders, including 275 OMIM diseases and 87 ORPHA syndromes, postulated to be emblematic of I/DSD conditions (see **Figure 2** and *Supplemental File 1*).

**Figure 2:**
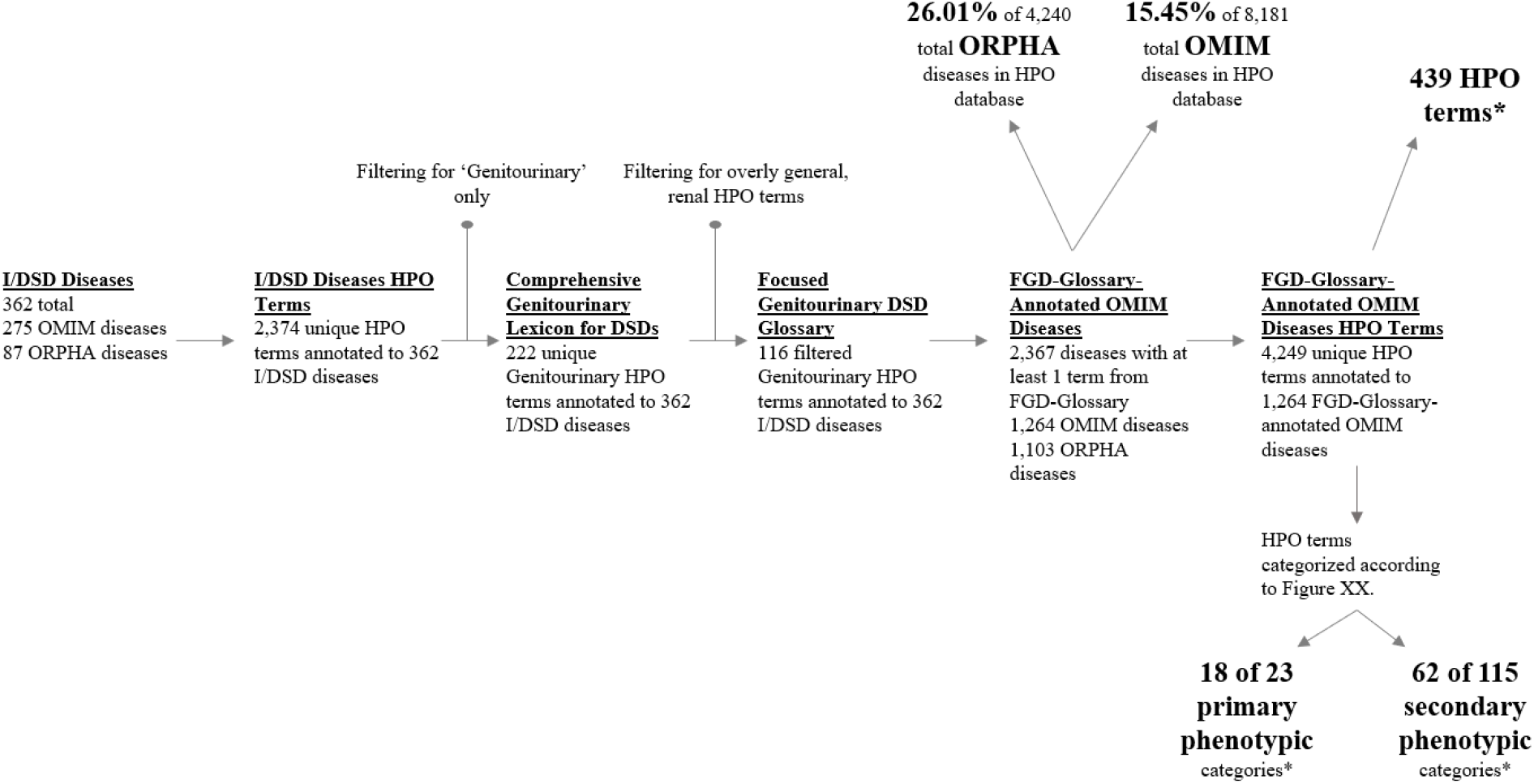
Overview of the Methodological Flow and Key Findings. This diagram provides a visual summary of the methodological process from the initial identification of FGD-Glossary-annotated OMIM diseases to the detailed analysis of HPO term distributions, highlighting the creation of the CGL-DSD and FGD-Glossary and the significant overlap of HPO terms across rare diseases. Differences in the frequency of HPO annotations across primary and secondary phenotypic categories are also depicted, illustrating the unique phenotypic landscape of I/DSD-related conditions. (*) denotes statistical significance with *p*-adj. < 0.05.

**Figure 3:**
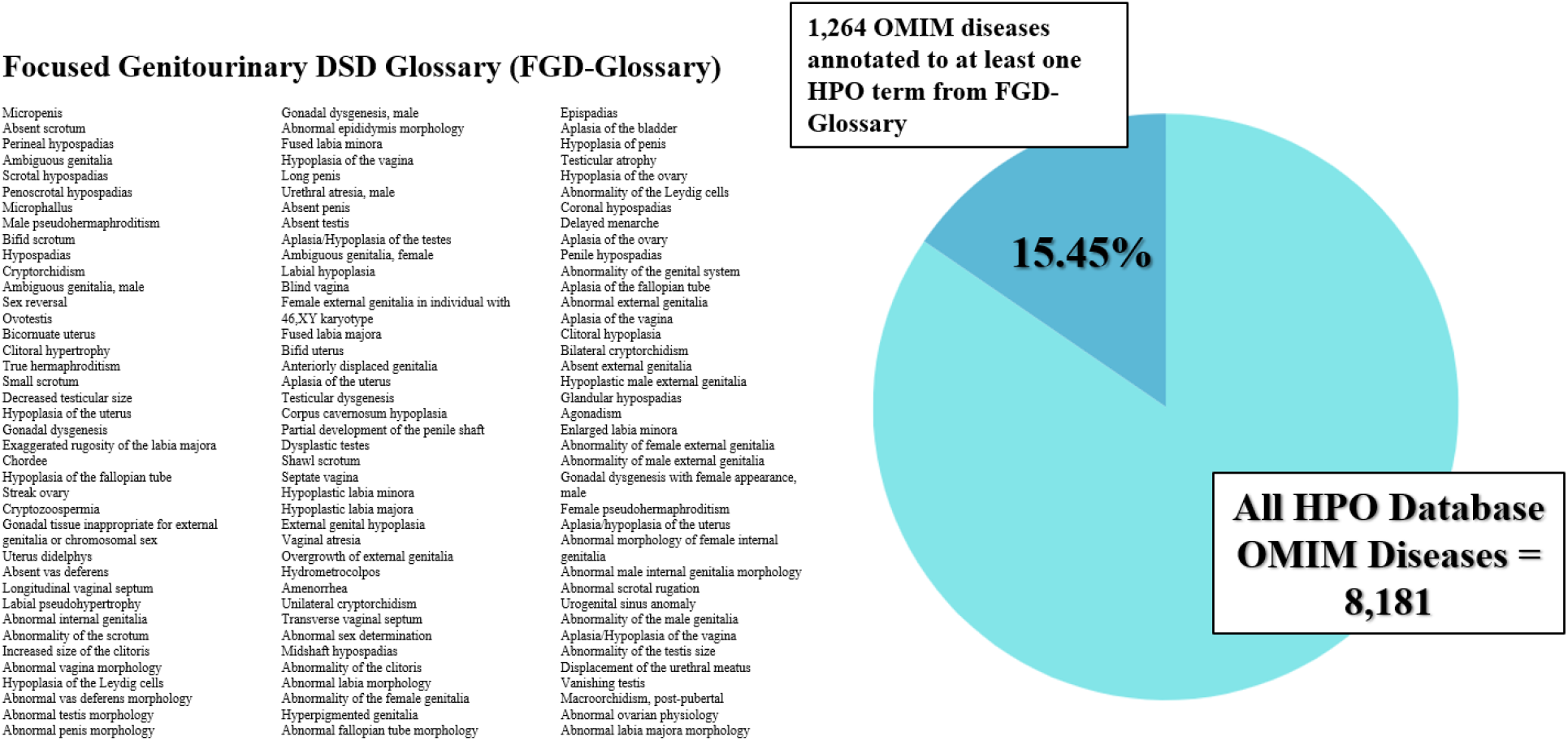
Distribution of FGD-Glossary HPO Terms Among OMIM Diseases. This pie chart illustrates that 15.5% of the 8,181 diseases cataloged in the HPO database include at least one term from the FGD-Glossary, highlighting the relevance of these terms in the broader context of genetic disorders.

### Formulation of Extensive HPO Lexicons

2,374 unique HPO terms were annotated to our list of I/DSD OMIM diseases and ORPHA syndromes. Filtering the list for ‘Genitourinary’ HPO terms resulted in the creation of the CGL-DSD, containing 222 HPO terms, that was then refined to the more FGD-Glossary encapsulating 116 HPO terms (see **Figure 2** and *Supplemental File 2*).

### Prevalence and Overlap of I/DSD Characteristics in HPO-Annotated Genetic Disorders

We identified 2,367 disorders, encompassing 1,264 OMIM diseases and 1,103 ORPHA syndromes, annotated to the HPO terms from the FGD-Glossary; further, we found 4,349 unique HPO terms annotated to the 1,264 FGD-Glossary-annotated OMIM diseases. This discovery implies a noteworthy overlap between a substantial proportion of the phenotypically described diseases in the HPO database and the HPO terms from our FGD-Glossary. Specifically, 15.5% of the 8,181 total HPO-annotated OMIM diseases and 26.0% of the 4,240 total HPO-annotated ORPHA syndromes were found to possess at least one HPO term from the FGD-Glossary. Considering the total of approximately 6,089 ORPHA syndromes, our findings reveal that 17.9% of these rare diseases are associated with an I/DSD HPO term. This significant percentage highlights the pervasiveness of I/DSD characteristics across a wide array of rare diseases (see **Figures 2** and **3**; *Supplemental File 3*).

### Assessing Differences in HPO Term Frequencies and Categorizations in FGD-Glossary-Annotated OMIM Diseases Using Z-Tests

#### Modes of Inheritance HPO Terms

Upon analyzing the HPO Modes of Inheritance terms, we found that 18 terms were represented in both the FGD-Glossary-annotated OMIM diseases and the entire HPO dataset. After conducting the proportions z-test and adjusting for multiple comparisons using the Benjamini-Hochberg procedure, six of these terms showed a statistically significant (*p*-adj.<0.05) difference in distribution. X-linked recessive inheritance (HP:0001419, *p*-adj.=5.7*10^-6^), sporadic inheritance (HP:0003745, *p*-adj.=5.1*10^-5^), autosomal recessive inheritance (HP:0000007, *p*-adj.=5.3*10^-3^), Y-linked inheritance (HP:0001450, *p*-adj.=1.4*10^-2^), and sex-limited expression (HP:0001470, *p*-adj.=4.0*10^-2^) had higher proportions in FGD-Glossary-annotated OMIM diseases, while autosomal dominant inheritance’s proportion (HP:0000006, *p*-adj.=1.4*10^-8^) was decreased. These terms are displayed in **Figure 4**.

**Figure 4:**
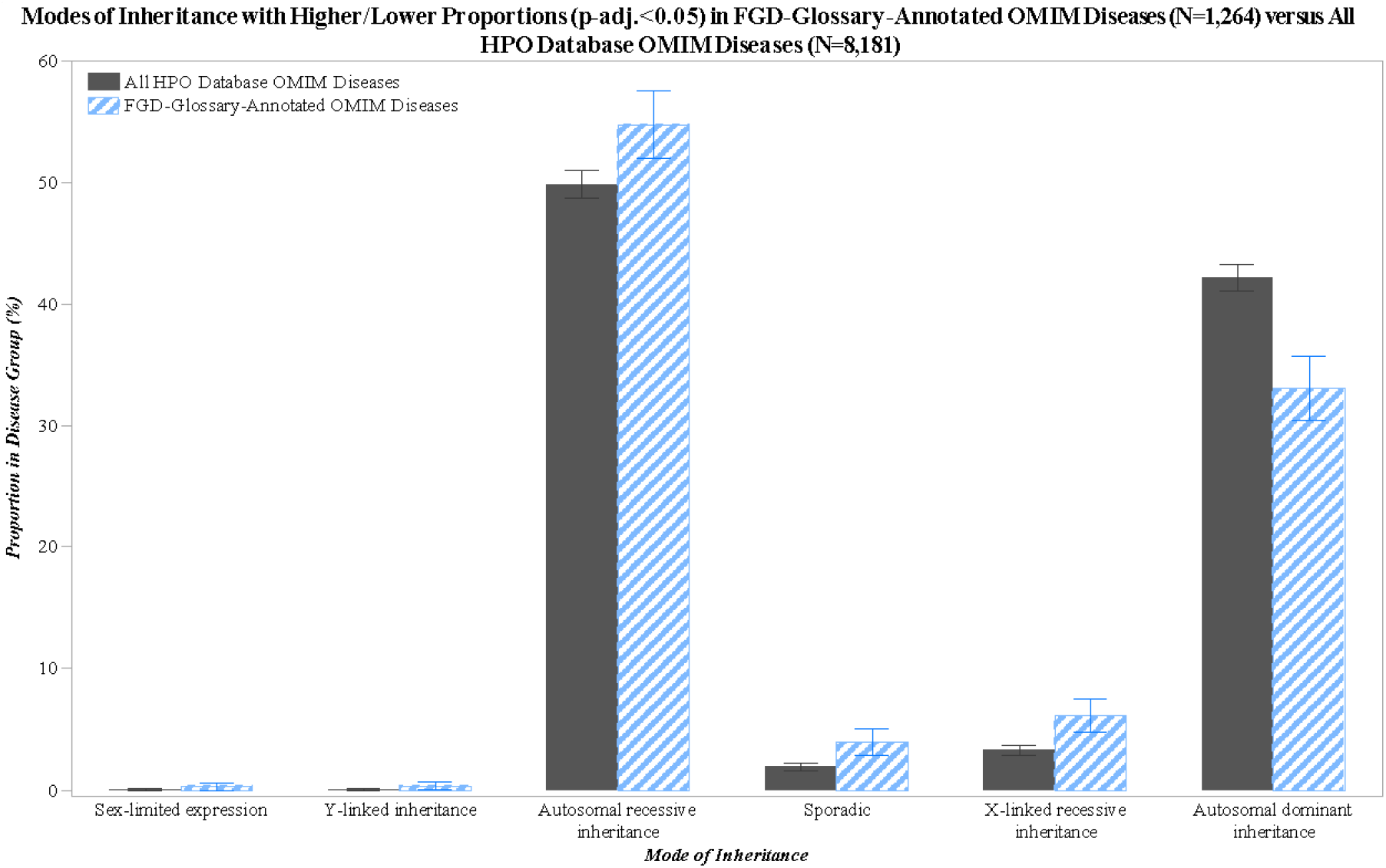
Comparative Distribution of HPO Modes of Inheritance in FGD-Glossary-Annotated OMIM Diseases. The bar graph highlights the six inheritance patterns with statistically significant differences in frequency between FGD-Glossary-annotated OMIM diseases and the entire HPO dataset, as determined by proportions z-tests with Benjamini-Hochberg p-value adjustments. Each error bar is constructed using ±2SE.

#### All HPO Terms

Our analysis further revealed that out of 4,349 unique HPO terms annotated to the OMIM diseases in our study (excluding FGD-Glossary HPO terms), 449 were found to have a statistically significant (*p*-adj.<0.05) difference in their frequency distribution. Among these, 439 HPO terms were more frequent than expected, while 10 were less frequent. These findings are summarized in Table 1, which provides an overview of the top 25 HPO terms with significant deviations in frequency.

**Table 1:** Significant Variations in HPO Term Frequencies in FGD-Glossary-Annotated OMIM Diseases. This table showcases the top 25 HPO terms with notable deviations in frequency compared to expected values, highlighting both the more frequently and less frequently occurring terms within the study’s OMIM diseases dataset.

| HPO | Name | Z-statistic | P-adj. | Direction |
| --- | --- | --- | --- | --- |
| HP:0000369 | Low-set ears | 17.9 | $9.2 \times 10^{-68}$ | Higher |
| HP:0003251 | Male infertility | 16.1 | $2.8 \times 10^{-55}$ | Higher |
| HP:0000316 | Hypertelorism | 15.1 | $1.4 \times 10^{-48}$ | Higher |
| HP:0000347 | Micrognathia | 14.9 | $2.2 \times 10^{-47}$ | Higher |
| HP:0001629 | Ventricular septal defect | 14.2 | $1.3 \times 10^{-42}$ | Higher |
| HP:0000786 | Primary amenorrhea | 13.0 | $8.1 \times 10^{-36}$ | Higher |
| HP:0000175 | Cleft palate | 13.0 | $9.3 \times 10^{-36}$ | Higher |
| HP:0000135 | Hypogonadism | 12.5 | $2.4 \times 10^{-33}$ | Higher |
| HP:0001631 | Atrial septal defect | 12.5 | $5.3 \times 10^{-33}$ | Higher |
| HP:0000358 | Posteriorly rotated ears | 12.4 | $7.2 \times 10^{-33}$ | Higher |
| HP:0004322 | Short stature | 12.2 | $1.2 \times 10^{-31}$ | Higher |
| HP:0000494 | Downslanted palpebral fissures | 12.2 | $1.2 \times 10^{-31}$ | Higher |
| HP:0000286 | Epicanthus | 12.0 | $1.8 \times 10^{-30}$ | Higher |
| HP:0000431 | Wide nasal bridge | 12.0 | $2.0 \times 10^{-30}$ | Higher |
| HP:0000218 | High palate | 11.9 | $5.2 \times 10^{-30}$ | Higher |
| HP:0001643 | Patent ductus arteriosus | 11.9 | $5.5 \times 10^{-30}$ | Higher |
| HP:0001511 | Intrauterine growth retardation | 11.8 | $1.6 \times 10^{-29}$ | Higher |
| HP:0000023 | Inguinal hernia | 11.2 | $8.2 \times 10^{-27}$ | Higher |
| HP:0001249 | Intellectual disability | 11.2 | $1.3 \times 10^{-26}$ | Higher |
| HP:0000470 | Short neck | 11.1 | $2.8 \times 10^{-26}$ | Higher |
| HP:0000252 | Microcephaly | 11.0 | $1.0 \times 10^{-25}$ | Higher |
| HP:0000463 | Anteverted nares | 10.7 | $1.2 \times 10^{-24}$ | Higher |
| HP:0005280 | Depressed nasal bridge | 10.6 | $4.5 \times 10^{-24}$ | Higher |
| HP:0000044 | Hypogonadotropic hypogonadism | 10.6 | $5.8 \times 10^{-24}$ | Higher |
| HP:0000219 | Thin upper lip vermillion | 10.5 | $1.3 \times 10^{-24}$ | Higher |

#### Primary Phenotypic Categories

For the primary phenotypic categories, our analysis included 22 categories with overlapping annotations between the FGD-Glossary-annotated and entire OMIM disease sets. The proportions z-test, followed by the Benjamini-Hochberg p-value adjustment, identified 18 categories with significant differences (*p*-adj.<0.05). These results indicate a notable variation in the representation of primary phenotypic categories between the select set of diseases and the comprehensive OMIM dataset. These distributions are represented in **Table 2** and **Figure 5**.

**Table 2:** Statistically Significant Differences in Primary Phenotypic Category Annotations. This table presents the 18 primary phenotypic categories that demonstrated statistically significant differences (*p*-adj.<0.05) in annotations when comparing the FGD-Glossary-annotated OMIM diseases with the entire set of OMIM diseases in the HPO database.

| Primary Phenotypic Category | Z-statistic | P-adj. | Direction |
| --- | --- | --- | --- |
| Abnormality of the genitourinary system | 38.3 | 0 | Higher |
| Abnormality of the nervous system | -29.8 | $1.12 \times 10^{-194}$ | Lower |
| Abnormality of head or neck | 27.4 | $1.48 \times 10^{-164}$ | Higher |
| Abnormality of metabolism/homeostasis | -19.6 | $7.15 \times 10^{-85}$ | Lower |
| Abnormality of the immune system | -14.7 | $3.79 \times 10^{-48}$ | Lower |
| Abnormality of the endocrine system | 13.7 | $2.24 \times 10^{-42}$ | Higher |
| Abnormality of the eye | -10.3 | $1.74 \times 10^{-24}$ | Lower |
| Abnormality of blood and blood-forming tissues | -9.6 | $1.52 \times 10^{-21}$ | Lower |
| Abnormality of the ear | 8.9 | $1.10 \times 10^{-18}$ | Higher |
| Constitutional symptom | -8.8 | $2.57 \times 10^{-18}$ | Lower |
| Abnormality of the breast | 8.1 | $1.27 \times 10^{-15}$ | Higher |
| Abnormal cellular phenotype | -7.8 | $8.18 \times 10^{-15}$ | Lower |
| Growth abnormality | 5.1 | $5.00 \times 10^{-7}$ | Higher |
| Abnormality of prenatal development or birth | 5.0 | $9.15 \times 10^{-7}$ | Higher |
| Abnormality of limbs | 4.6 | $6.53 \times 10^{-6}$ | Higher |
| Abnormality of the cardiovascular system | -4.1 | $5.09 \times 10^{-5}$ | Lower |
| Neoplasm | 2.6 | 0.0124 | Higher |
| Abnormality of the digestive system | 2.2 | 0.0357 | Higher |

**Figure 5:**
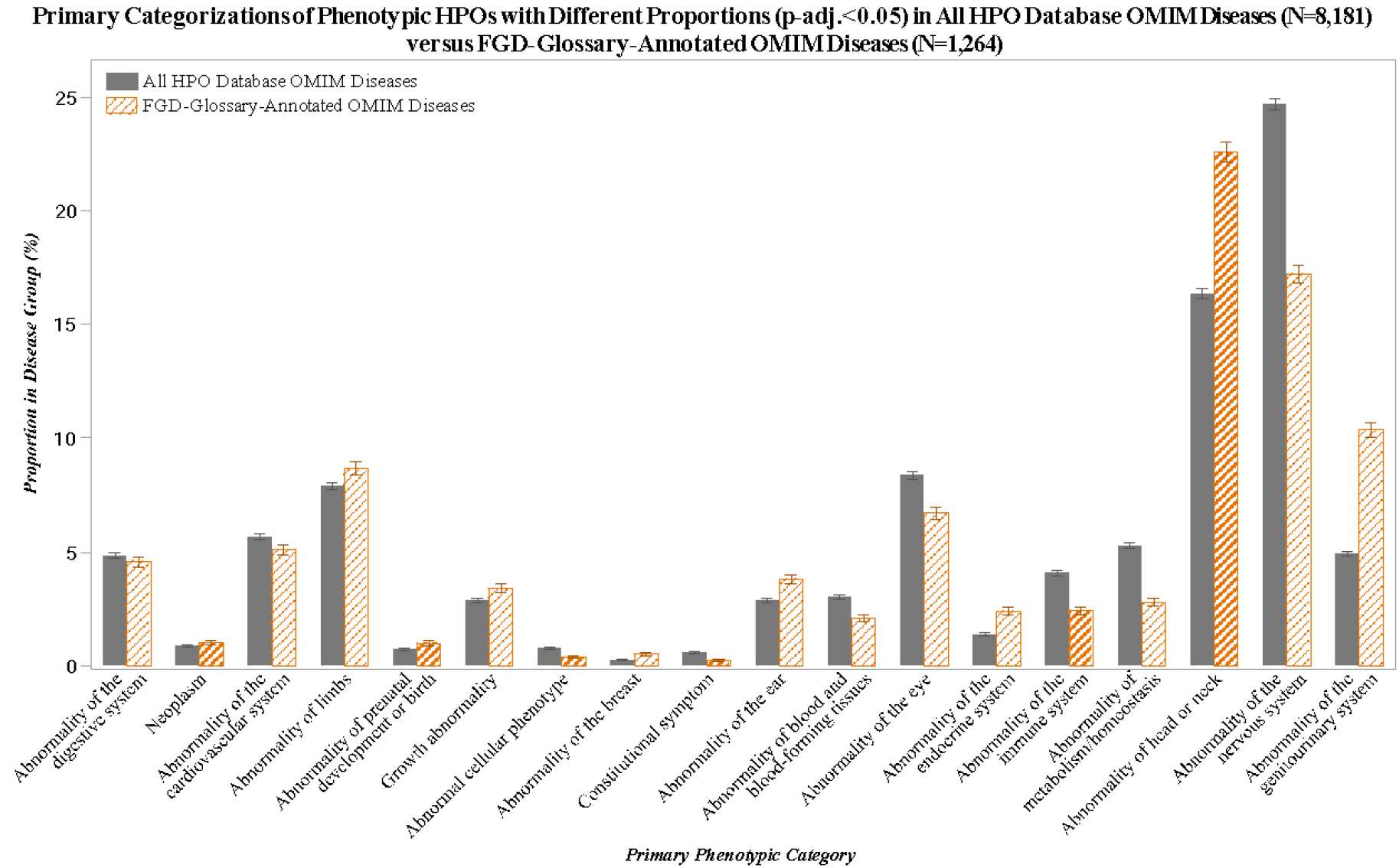
Prevalence of Primary Phenotypic Categories. This bar chart contrasts the occurrence of 18 primary phenotypic categories (*p*-adj.<0.05) between FGD-Glossary-annotated OMIM diseases and the full HPO database OMIM dataset. Each error bar is constructed using ±2SE.

The “Abnormality of Head or Neck’’ and “Abnormality of the Endocrine System” primary phenotypic categories were further analyzed to identify the individual HPO terms that exhibited the most pronounced frequency differences within the FGD-Glossary-annotated OMIM diseases. For the “Abnormality of Head or Neck’’ category, the following HPO terms were significantly more frequent in the FGD-Glossary-annotated diseases: micrognathia (HP:0000347, *p*-adj.=1.18*10^-47^), cleft palate (HP:0000175, *p*-adj.=4.35*10^-36^), downslanted palpebral fissures (HP:0000494, *p*-adj.=6.56*10^-32^), epicanthus (HP:0000286, *p*-adj.=7.34*10^-31^), wide nasal bridge (HP:0000431, *p*-adj.=7.34*10^-31^), high palate (HP:0000218, *p*-adj.=1.76*10^-30^), and short neck (HP:0000470, *p*-adj.=1.09*10^-26^), among others. In the “Abnormality of the Endocrine System” category, HPO terms such as hypogonadism (HP:0000135, *p*-adj.=7.16*10^-34^), hypogonadotropic hypogonadism (HP:0000044, *p*-adj.=2.56*10^-24^), and hypergonadotropic hypogonadism (HP:0000815, *p*-adj.=2.15*10^-21^) were notably more prevalent in the FGD-Glossary subset. The top 15 HPO terms for each category are visually represented in **Figure 6**.

**Figure 6:**
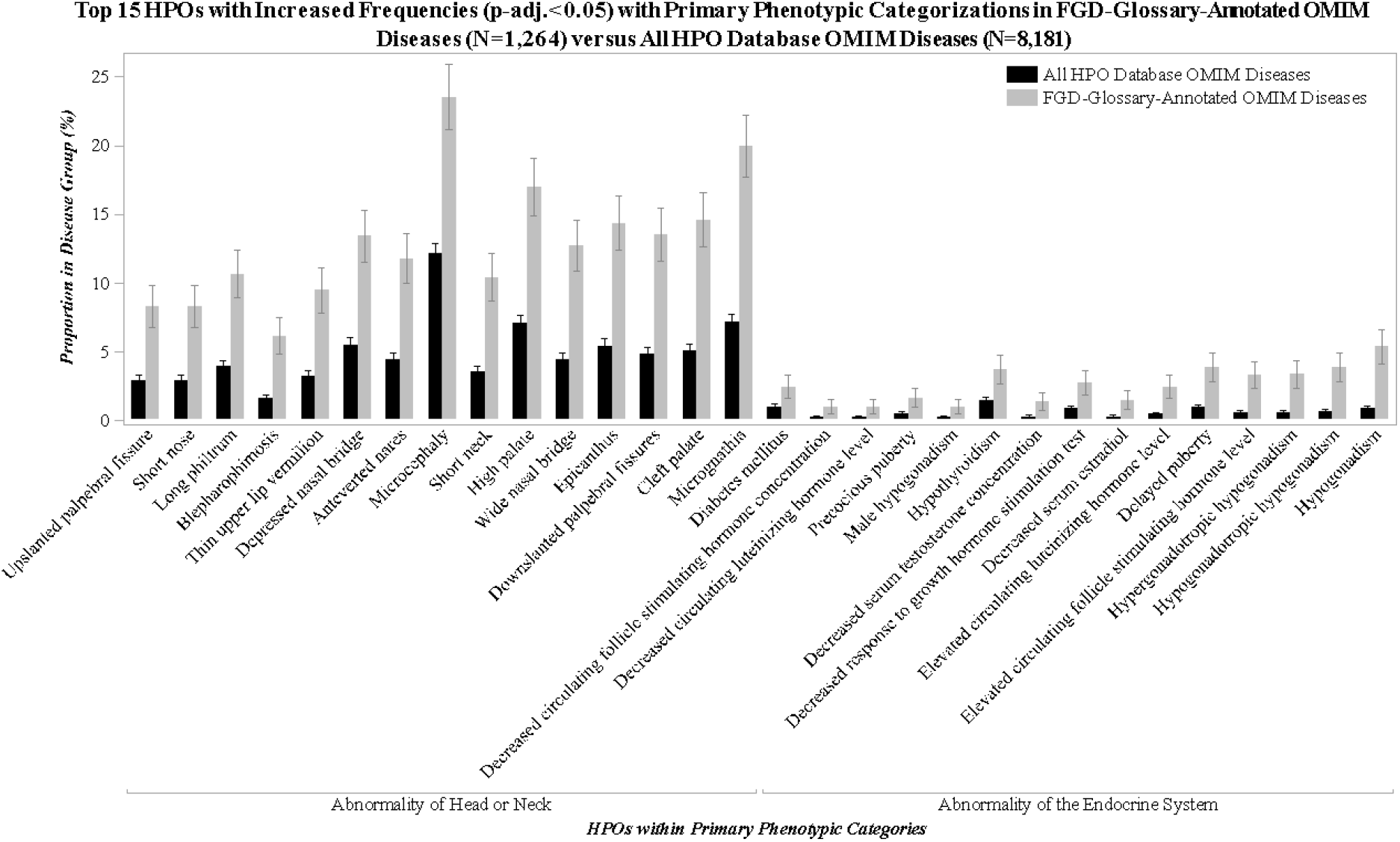
Top 15 Differentially Represented HPO Terms in “Abnormality of Head or Neck” and “Abnormality of the Endocrine System” Primary Phenotypic Categories. This figure displays a bar chart of the 15 HPO terms with the most significant differences in frequency (*p*-adj.<0.05) between FGD-Glossary-annotated OMIM diseases and the entire HPO database OMIM dataset for each category. Each error bar is constructed using ±2SE.

#### Secondary Phenotypic Categories

In the secondary phenotypic categories, we examined 114 categories that were common to both the select and entire sets of OMIM diseases. The statistical analysis revealed that 62 of these categories showed significant differences in HPO annotation proportions after adjusting the *p*-values (*p*-adj.<0.05). This highlights the disparities in secondary phenotypic category representation between the select group of FGD-Glossary-annotated OMIM diseases and the entire HPO database OMIM disease collection. The detailed distributions for the statistically significant (*p*-adj.<0.05) increased (N=31) and decreased (N=31) secondary categories are depicted in **Figures 7** and **8**, respectively.

**Figure 7:**
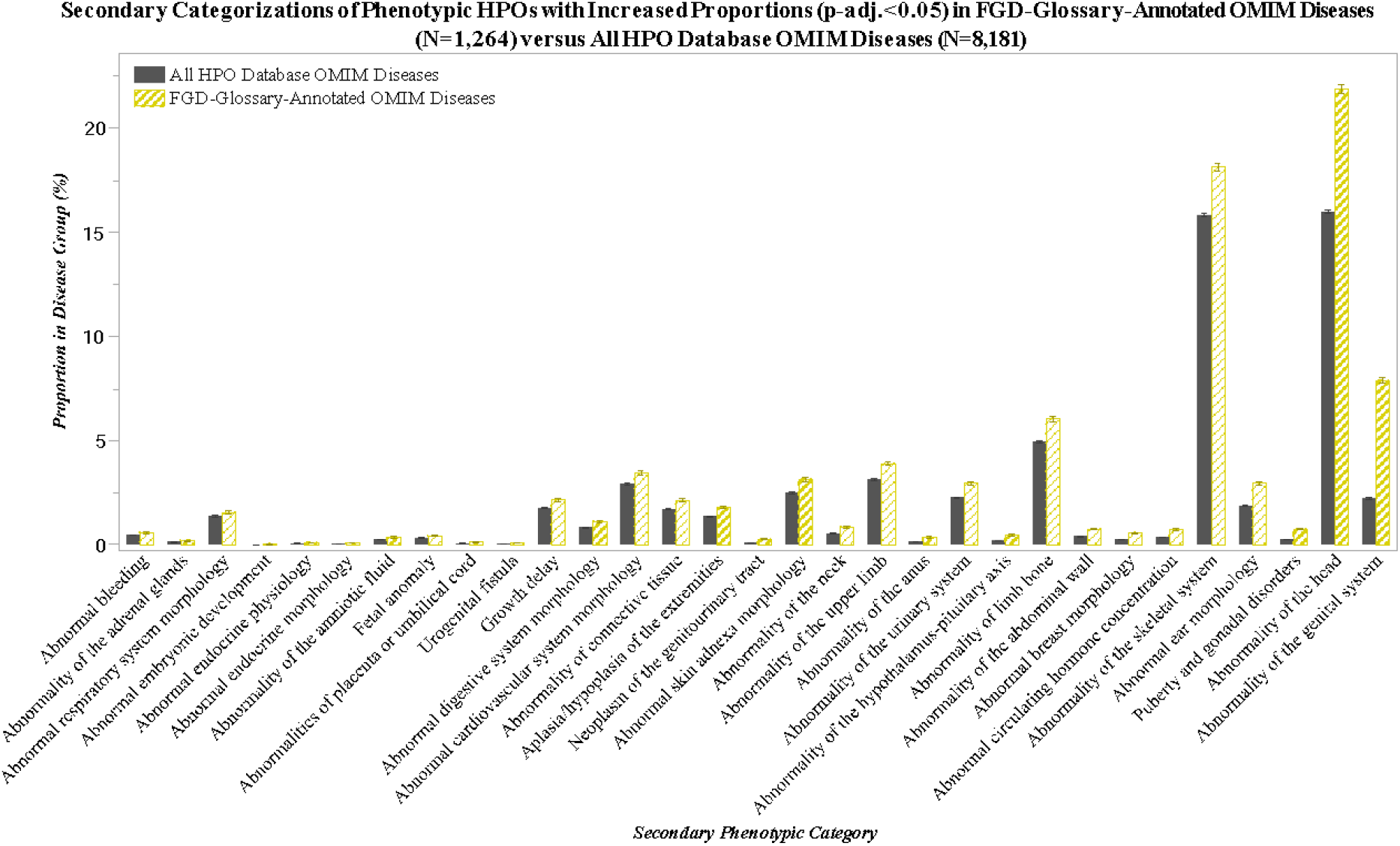
Distribution of Increased Secondary Phenotypic Categories. This figure illustrates the 31 secondary phenotypic categories with a statistically significant increase in HPO annotation proportions among the select group of FGD-Glossary-annotated OMIM diseases compared to the entire HPO database OMIM disease collection (*p*-adj.<0.05). The distribution highlights the specific phenotypic patterns that are more prevalent in the select group. Each error bar is constructed using ±2SE.

**Figure 8:**
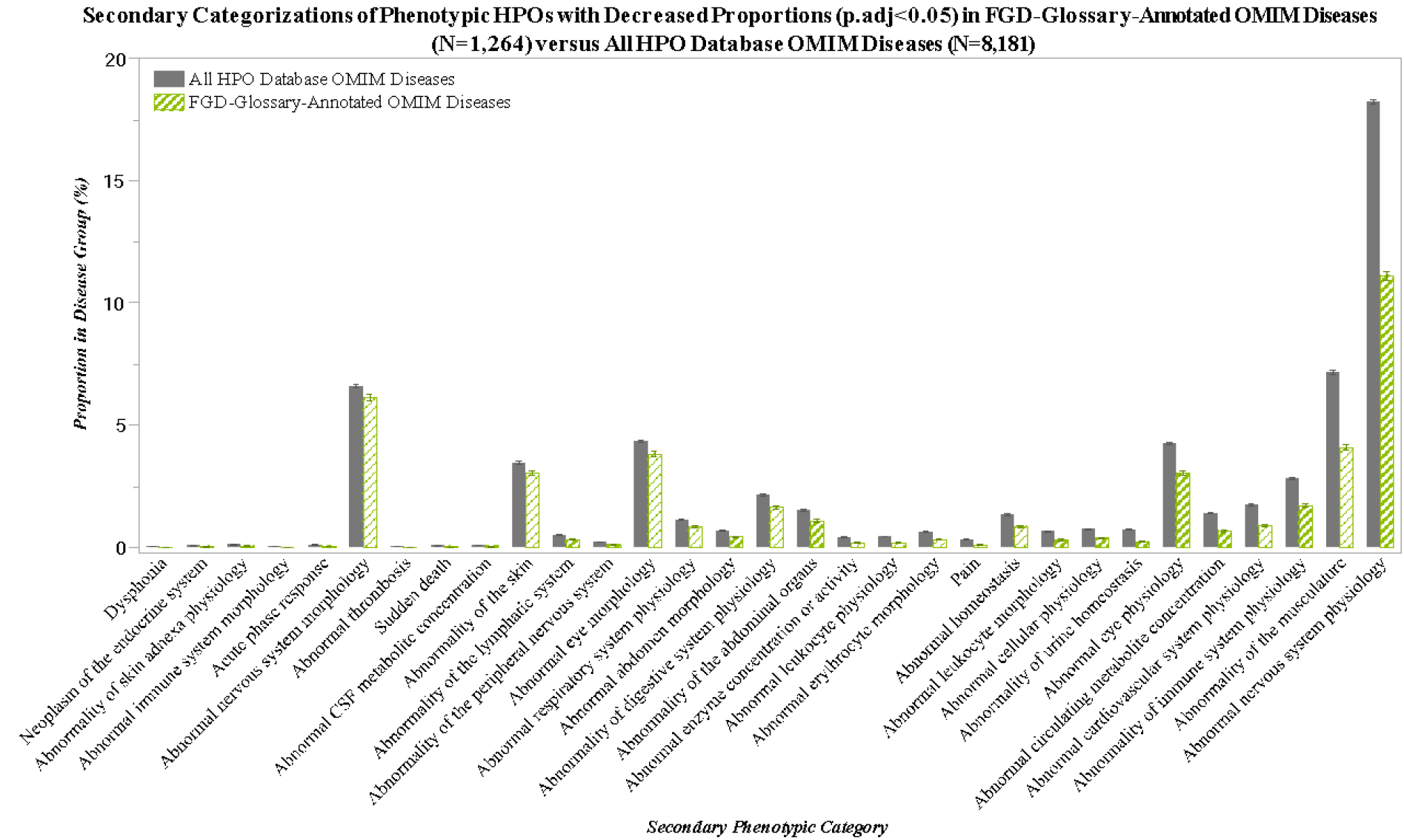
Distribution of Decreased Secondary Phenotypic Categories. This figure depicts the 31 secondary phenotypic categories with a statistically significant decrease in HPO annotation proportions in the select group of FGD-Glossary-annotated OMIM diseases relative to the entire HPO database OMIM disease collection (*p*-adj.<0.05). The distribution highlights the phenotypic categories that are underrepresented in the select group. Each error bar is constructed using ±2SE.

In a focused analysis of the top five increased secondary phenotypic categories, individual HPO terms that were significantly more prevalent within these categories were identified. For the “Abnormality of the Head’’ category, the three HPO terms with the highest significance of increased frequency were micrognathia (HP:0000347, *p*-adj.=1.14*10^-47^), cleft palate (HP:0000175, *p*-adj.=4.21*10^-36^), and downslanted palpebral fissures (HP:0000494, *p*-adj.=6.35*10^-32^). In the “Puberty and Gonadal Disorders” category, hypogonadism (HP:0000135, *p*-adj.=1.00*10^-34^), hypogonadotropic hypogonadism (HP:0000044, *p*-adj.=3.58*10^-25^), and hypergonadotropic hypogonadism (HP:0000815, *p*-adj.=3.01*10^-22^) were the top three HPO terms exhibiting increased prevalence. Regarding “Abnormal Ear Morphology,” the most increased HPO terms were low-set ears (HP:0000369, *p*-adj.=1.61*10^-69^), posteriorly rotated ears (HP:0000358, *p*-adj.=6.26*10^-34^), and macrotia (HP:0000400, *p*-adj.=3.81*10^-17^). For “Abnormality of the Skeletal System,” the top three HPO terms were micrognathia (HP:0000347, *p*-adj.=1.81*10^-47^), short neck (HP:0000470, *p*-adj.=5.85*10^-26^), and microcephaly (HP:0000252, *p*-adj.=1.47*10^-25^). Lastly, within the “Abnormal Circulating Hormone Concentration” category, the three most increased HPO terms were elevated circulating follicle stimulating hormone level (HP:0008232, *p*-adj.=5.47*10^-20^), elevated circulating luteinizing hormone level (HP:0011969, *p*-adj.=1.28*10^-14^), and decreased serum estradiol (HP:0008214, *p*-adj.=1.89*10^-9^). The top six HPOs within these secondary phenotypic categories are displayed in **Figure 9**.

**Figure 9:**
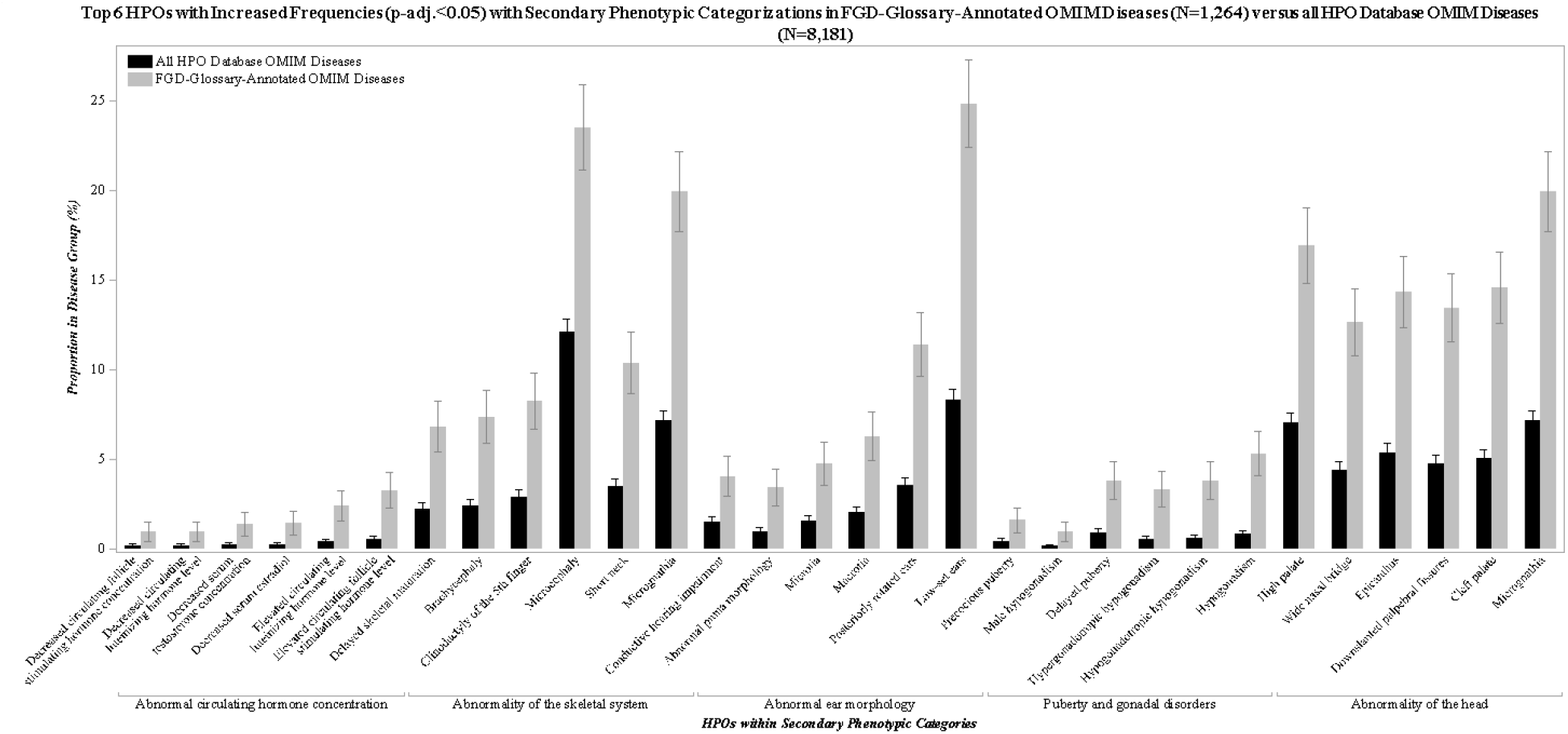
Top HPO Terms in Increased Secondary Phenotypic Categories. This graph presents the six HPO terms with the highest increase in frequency within each of the five secondary phenotypic categories identified as significantly elevated in FGD-Glossary-annotated OMIM diseases. Categories include Abnormality of the Head, Puberty and Gonadal Disorders, Abnormal Ear Morphology, Abnormality of the Skeletal System, and Abnormal Circulating Hormone Concentration. Each error bar is constructed using ±2SE.

## Discussion

### Systemic Implications of I/DSD Traits Necessitate a Multifaceted Approach to Diagnosis and Management

The 15.5% overlap of FGD-Glossary-annotated OMIM diseases with all HPO database OMIM diseases suggests that a substantial portion of genetic disorders cataloged in OMIM exhibit traits related to I/DSD conditions. This finding implies that I/DSD traits may not be as isolated as previously thought and could be indicative of broader genetic patterns or syndromes. The broader implications of I/DSD traits on other systems like the genitourinary, endocrine, and craniofacial regions highlight the interconnectedness of developmental pathways. Indeed, the critical signaling pathways involved in organogenesis, such as WNT, NOTCH, TGFβ, HH, and various RTKs, are also pivotal during sex determination and gonad development, underscoring a shared molecular framework that governs the development of diverse organ systems (Windley & Wilhelm, 2016). This commonality points to the potential for cross-system insights and the need for an integrated understanding of developmental biology.

I/DSD traits can have systemic implications, affecting multiple organ systems. This aligns with the current understanding that sex development is a complex, multi-system process. It has been previously identified that over a quarter of DSD cases had an additional condition, such as cardiac anomalies or central nervous system disorders, with a significant prevalence of 27%, which is over 10 times the birth prevalence of congenital anomalies (Cox et al., 2014). The complexity of these conditions is further compounded by the recognition that DSD may be caused by oligogenic variants working in concert, as a single variant cannot explain the phenotype, suggesting a need for comprehensive genetic analysis approaches, such as whole exome sequencing (WES) and whole genome sequencing (WGS), despite the challenges in their interpretation (Kouri, Sommer, & Flück, 2023). To ensure a comprehensive approach, a detailed medical history, physical examination, hormone measurements, and genetic testing are emphasized as crucial for the accurate diagnosis of individuals with DSDs, underscoring the complex nature of sex development as a multi-system process (Lee et al., 2016).

Further, this research highlights the wide range of presentations associated with I/DSD and the importance for clinicians to recognize this diversity. A limited perception of I/DSD phenotypes can result in incorrect or delayed diagnoses, directly impacting patient care. The overlap of physical traits and hormonal profiles among various DSD conditions further complicates accurate diagnosis, making it one of the most demanding tasks in healthcare (Cools et al., 2018). It is imperative that medical professionals are educated about the full spectrum of I/DSD manifestations, which can range from minor hormonal discrepancies to conspicuous physical differences. For instance, congenital adrenal hyperplasia can lead to ambiguous genitalia in newborn girls due to excessive androgens, whereas boys with complete androgen insensitivity syndrome may appear typically female, despite having a 46, XY chromosome set (Pasterski, Prentice, & Hughes, 2010). These cases exemplify the extensive variability within I/DSD conditions, underscoring the necessity for a detailed and discerning approach to both diagnosis and management.

Finally, we observed a notable decrease in the frequency of HPO terms related to nervous system abnormalities among FGD-Glossary-annotated OMIM diseases. This finding presents a contrast to the previous research, which indicates an increased prevalence of cognitive and motor disturbances and psychiatric conditions in individuals with I/DSD, suggesting that such nervous system involvements are clinically significant in this patient population (Cools et al., 2018; Cox et al., 2014). The discrepancy may reflect a limitation in the HPO’s ability to capture non-observable clinical findings as effectively as it does observable phenotypic traits. For instance, the increased prevalence of head, ear, and eye deformities in FGD-annotated OMIM diseases implies that HPO terms are more reflective of readily apparent physical manifestations. This highlights the necessity for ongoing enhancement of HPO characterizations to ensure thorough and nuanced documentation of both overt and subtle clinical findings. The comprehensive reporting of these observations is critical for advancing our understanding of DSDs and improving patient care.

## Supporting information

Supplemental File 4_Categorizations and Z-Tests

Supplemental File 2_Focused Genitourinary DSD Glossary

Supplemental File 1_I-DSD Diseases HPO Annotations

Supplemental File 3_HPOs Annotated to FGD-Glossary-Annotated OMIM

## Data Availability

All data produced in the present study are available upon reasonable request to the authors.

